# Rationale, design, analysis plan and baseline characteristics of a cluster randomized controlled trial for management of salt intake and salt procurement in hypertensive individuals and households in rural Central India through a Community Health Worker delivered Behavior Change Communication Intervention

**DOI:** 10.1101/2023.10.13.23297011

**Authors:** Raunaq Singh Nagi, Sanjeev Kumar, Pankaj Prasad

**Author notes:** **Corresponding author:** Raunaq Singh Nagi Graduate Student (PhD), Department of Community & Family Medicine, All India Institute of Medical Sciences, Bhopal--462020 (MP), India. **Email addresses:**. **Author contributions:** RSN, SK and PP contributed to conceptualization of the study and preparation of study tools, SK and PP were involved in supervision and obtaining permissions for the study, RSN was responsible for data collection and analysis. All the authors contributed to reviewing, editing, and approving the final manuscript.

## Abstract

**Background:** Cardiovascular diseases (CVD) contribute to highest number of non-communicable diseases associated morbidity and mortality. Uncontrolled hypertension has been linked to development and progression of CVDs. The global age-standardized prevalence of hypertension in 30-79 years age group is 32% for women and 34% for men. Hypertension is a metabolic risk factor that is drastically affected by dietary salt. Excessive dietary salt consumption is a behavioral risk factor and has been a WHO global target for hypertension and CVD management. The per capita global and national dietary salt consumption estimates exceed the recommended cut-off of 5g/day. Apart from policy changes, dietary salt consumption reduction through health promotion activities has been mandated by WHO and initiated in many countries. Community health workers (CHWs) have been identified as a target cadre for successful implementation of health promotion interventions.

We aim to ascertain the effectiveness of various counseling modes on relative reduction of per capita dietary salt intake and household salt procurement.

**Methods:** An open label, factorial, cluster-RCT was executed in rural central India. Intervention arms were individual, group and mixed (group followed by individual) counseling and one standard care arm.

**Results:** A total of 127 adult hypertensive individuals were recruited in the study. One CHW was considered as one cluster and a total of eight (8) clusters were randomized for the study. The baseline characteristics of the participants have been presented.

**Conclusion:** The results of this trial will inform up on effectiveness of different modes of behavior change communication intervention on reducing dietary salt-intake and procurement.

## 1. Background

Treatment of hypertension in India is unsatisfactory and requires further strengthening of primary healthcare system.[1] As per pooled estimates of studies from 2016-2020, only 22.5% hypertensive Indians have their blood pressure under control.[2] It is estimated that 36.5% hypertensive individuals are lost at screening stage itself.[3] Raised blood pressure is an important risk factor for cardiovascular diseases (CVDs).[4] In 2016, CVDs accounted for 28.1% total annual deaths and 14.1% of total annual DALYs, with Ischemic heart Disease (IHD; 8.7% DALYs; 17.8% deaths) and stroke (3.5% DALYs; 7.1% deaths) as leading CVDs.[5] Raised blood pressure is a direct outcome of excessive salt intake. High salt (sodium) intake leads to disruption of metabolic balance of sodium and thus, to fluid retention.[6] Global Burden of Disease Study Diet Collaborators report high intake of sodium to be foremost dietary risk factor, with 3 million deaths and 70 million Disability-Adjusted Life Years (DALYs) globally in 2017.[7] The age-standardized death rates due to high sodium intake were reported to be 32.96 per 100000 in males and 15.71 per 100000 in females, in the year 2019.[8] The World Health Organization (WHO) recommends reduction of 30% mean population level sodium intake by 2025 and below 2000mg/day beyond 2025, via multitude of strategies such as, promotion of sodium reduced salt, integration of community-level behavioral health promotion activities targeting dietary salt in primary care, front-of-package labeling, salt taxation, and others. More than 90 national level salt reduction initiatives have been launched by WHO member states, most of which are multi-sectoral.[9] Community-based lifestyle modification interventions delivered by trained primary healthcare workers has been shown to significantly reduce systolic and diastolic blood pressure in rural India.[10] However, the cost of lifestyle modification interventions for CVD risk reduction is restricted in Low- and Middle-Income Countries (LMICs) due to financial constraints.[11] Pooled estimates of effectiveness of Community Health Workers (CHWs) led interventions in reducing systolic and diastolic blood pressures (SBP and DBP) in developing countries have been comparable to medically-trained healthcare worker led interventions..[12] Task-shifting to non-physician healthcare workers (such as CHWs) for patient education has emerged as a noteworthy implementation strategy for CVD management.[13]

Community-based behavior change communication interventions (BCCIs) have been successful in reducing risky behaviors in rural communities.[14] Perceived cultural similarity with CHWs has enabled participation and adoption of healthy behaviors via health communication.[15]Well-designed BCCIs have efficient and wider reach.[16]r Culturally-adopted community-based interventions targeting NCDs have been delivered by CHWs in rural India, and are feasible and scalable.[17] Two modes of intervention, group and individual are widely used to target NCD-associated unhealthy behaviors.[18,19] However, the ambiguity regarding effectiveness of these modes of intervention compared to one-another,[18,19] causes a dilemma for policymakers while allocating monetary and human resources.

### Rationale

Reduction of per capita salt (sodium) consumption in hypertensive individuals is a global target for controlling development of CVDs and CVD associated morbidity and mortality. National governments and international agencies have mounted responses to regulate discretionary use of salt. One of the most crucial approaches in this context is introduction of behavior change communication strategies under the umbrella of health promotion and patient education. Engagement of CHWs in health promotion and patient education can prove to be an effective task-shifting strategy and is considered appropriate due to cultural reasons.

We present the rationale, design, analysis plan, and baseline characteristics for a cluster-randomized controlled trial (RCT) for evaluation of effectiveness of a culturally adapted, consumer (hypertensive individual) informed, community-based behavior change communication intervention (BCCI) in reducing salt intake and procurement in rural central India.

## 2. Methods/Design

### 2.1. Study aims

The study was conceived to assess the effectiveness of standalone individual and group counseling BCCI modes in reducing per capita daily dietary salt intake in hypertensive individuals and in reducing household level monthly salt procurement; and to ascertain any interactive effect of both counseling modes; and to compare these with standard care.

### 2.2. Study design

This study was an open-label cluster randomized controlled trial of factorial design consisting of four arms. Accredited Social Health Activists (ASHA-a word that translates to ‘hope’ in Hindi) are female CHWs in rural India. As per guidelines of National Rural Health Mission (NRHM, 2005-06-when ASHA program was launched in the country) one ASHA is alloted per 1000 residents of a village.[20] Hence, one ASHA was considered as one cluster.

The trial was conducted in 2x2 factorial design where clusters were randomized into one of the four arms: Individual Counseling (IC), Group Counseling (GC), Mixed Counseling (MC) and Standard Care (SC).

### 2.3. Study setting

The study was conducted in villages under the *Goharganj* Community Health Center (CHC) in the *Obedullaganj* administrative block of *Raisen* district of Madhya Pradesh, India. Under the Indian Public Health Standard (IPHS) guidelines, CHCs are secondary healthcare units catering to a population of 50,000. And provide administrative and clinical support to primary healthcare units called Health and Wellness Centers (HWCs) that cater to a population of 5,000. These HWCs are direct reporting centers of ASHAs in rural and urban areas alike. We selected two HWCs-*Goharganj* and *Dhamdhusar*-within five km range of CHC-*Goharganj*.

### 2.4. Study participants and sample size

As mentioned earlier, we selected HWCs located within five km distance from the CHC. All the ASHAs in both the HWCs were approached for participation. They were briefed regarding the requirements of the study and intervention process and were requested to provide consent for participation.

*Obedullaganj* administrative block consists of a population of 22,845 individuals, of which 12,000 are males (Census, 2011). National Family Health Survey round five (NFHS-5) reports the prevalence of hypertension in rural men to be 21.5% and in rural women to be 19.9% in individuals above the age of 15 years.[21]

Sample size calculation was done using effect sizes for individual, group and interaction interventions, derived from the studies of *Yamasaki T* et al (2015)[22], *Hirota S* et al (2012)[23] and *Daivadanam M* et al (2018)[24], respectively. Calculation of effect sizes (Cohen’s F) was done using G*power[25] application. The effect sizes were found to be 0.174 for group, 0.85 for individual and 0.425 for mixed counselling. The sample size calculation for factorial design was conducted using *easypower* package[26] for R programming language[27] in RStudio IDE[28]. For a desired power of 80% and type I error probability of 0.05, the total sample size was achieved to be 42 (11 each arm), seven (two each arm) and 15 (four each arm), respectively for the three effects. Up on adjusting for design effect of 1.5 to account for variations due to cluster design and attrition rate of 10% a final sample size of 63 is attained. We also calculated the sample size for evaluation of within arm variation using G*power software. Studies by *Yamasaki T* et al and *Hirota S* et al (referred above) were used for calculation. T-test: means: difference between two dependent means (matched pairs) was used to calculate the sample sizes. Total sample size of 51 and 29 was attained for assessment of group and individual counselling respectively, at 80% power and alpha error probability of 5%.

Considering the largest sample size, to attain a sufficient power at least 51 participants shall be required to receive group counselling. Hence, we 26 participants were to be recruited in group counselling and mixed counselling arms each. Consequently, minimum 26 participants were required to be recruited for each arm (1:1:1:1 ratio for all the arms). A total sample size of 104 participants was required to be achieved.

### 2.5. Recruitment

Based on the total sample size a total of eight ASHAs were recruited, two in each of the arms. Hence, the size of each cluster was proposed to be 13 participants.

List of eligible participants was obtained from the records of NCD patients maintained at the HWC. All the individuals above the age of 30 years and with a systolic blood pressure ≥ 140 mm Hg and/or diastolic blood pressure ≥ 00 mm Hg (measured in-clinic by trained healthcare worker) were considered to be hypertensive[29] and were approached for participation in the study. Figure 1 shows the COSORT[30] flow diagram of the participants recruited in the study. Table 1 contains the eligibility criteria for clusters and trial participants.

**Figure 1.**
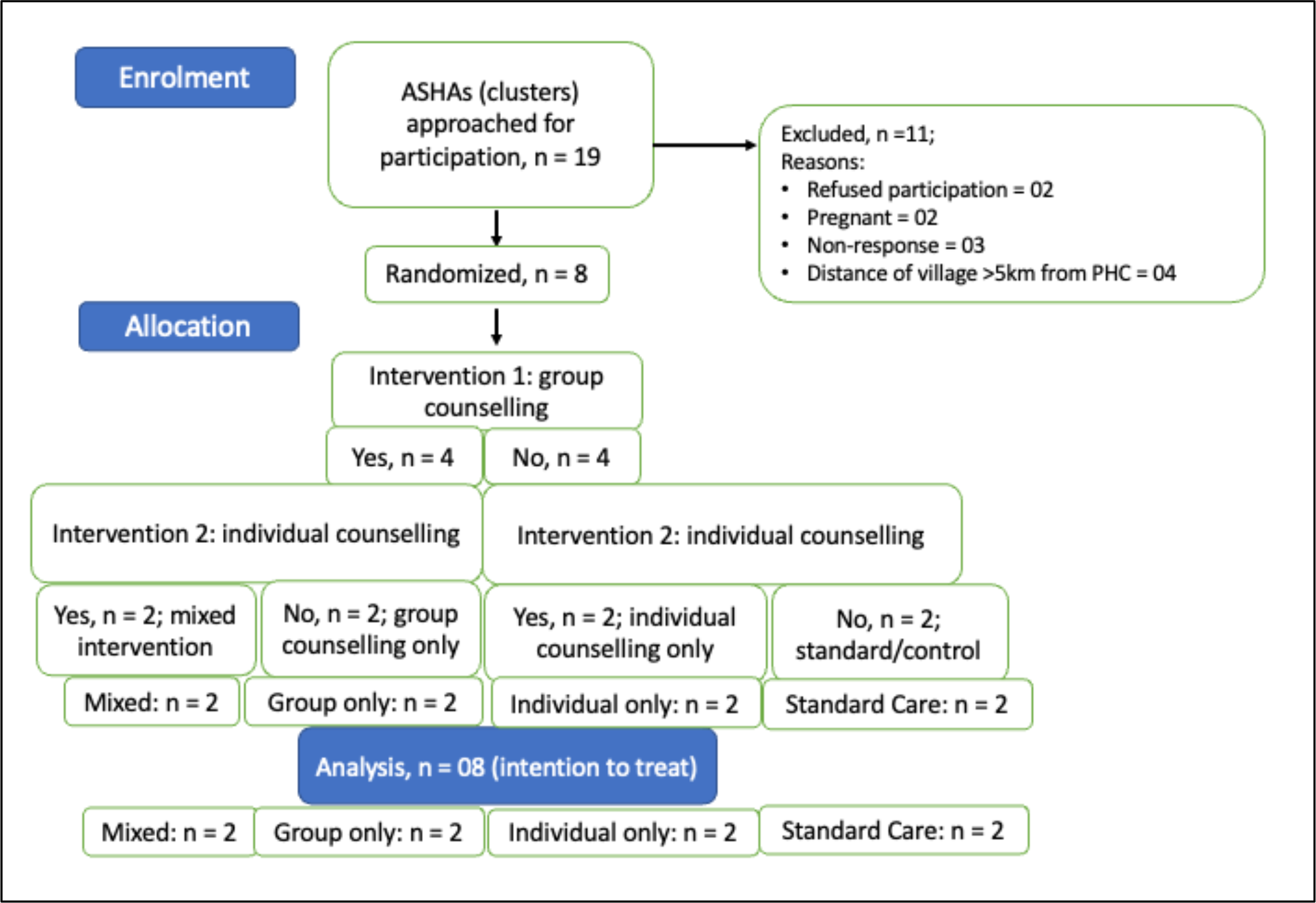
CONSORT flow diagram of the clusters of intervention.

**Table 1.** Eligibility criteria for clusters and participants.

| Unit | Inclusion | Exclusion |
| --- | --- | --- |
| Cluster/ASHA | Reports to HWC within 5 km periphery of CHC | Pregnant/lactating mothers<br>Non-response to meeting requests on two consecutive phone calls |
| Participants | At least 30 years of age<br>No comorbidity<br>Clinically confirmed case of HTN (SBP of 140 mm Hg or more and/or DBP of 90 mm Hg or more) | Pregnant/lactating mothers<br>Self-declared or clinically recorded physical or learning limitation which might impede attendance and/or comprehension of verbal instructions or pictorial data |

### 2.6. Randomization and allocation

Medical Officer (MO, medical doctor who oversees the function of their respective centers) of the CHC performed randomization and random allocation of the clusters to intervention arms. The MO assigned sequential numbers to ASHAs and concealed the number in optically opaque papers folded twice. Another staff member of the CHC prepared sequentially numbered opaque containers with codes for intervention arms. Folded papers with codes of ASHA were randomly placed in these boxes by the MO (two folded papers in each box). The MO was responsible for storage the boxes and the codes, that were provided to the investigators before start of the intervention.

### 2.7. Interventions

ASHAs have been tasked with community-level identification of individuals suffering from NCDs. Additionally, they are also responsible for conduction of health promotion activities and community mobilization for such activities. Furthermore, ASHAs are also a mediator for task-sharing activities for enhanced primary care coverage and proper referral. All interventions of this trial were executed by ASHAs after receiving relevant training to understand dietary salt-related variables viz, detrimental effects of excessive salt consumption, correct daily salt consumption limit, overt and covert sources of dietary salt, dietary practices that lead to excessive salt intake, and behavioral targets for salt consumption and procurement reduction. Quality control was ensured via post-training and mid-intervention assessment. The total duration of intervention was six months. An induction session was conducted before commencement of intervention. Delivery of intervention spanned over two months with incremental change in intervals between consecutive intervention sessions (from one week to one month).

Interventions consisted of messages in vernacular language that were enunciated verbatim by the ASHAs. These messages were accompanied by colorful pictorial references to be shown to the participants with respective message. The messages and pictorial references were printed on glossy paper and bound in the form of a flipchart. Types of messages included in the intervention have been provided in Supplementary Data.

Additionally, interventions were stage-matched according to readiness or stage of change of individual participant. Readiness to change was assessed during each session and customized intervention was provided to the participant. ASHAs assessed stage of change by using an assessment question, answers of which were printed in different colors. These colors were subsequently matched with similarly colored pages in the intervention modules and corresponding messages were delivered to the participants

#### 2.7.1. Individual counseling

The messages/verbatims for IC were grouped according to stage of change of the participant. ASHAs assessed the stage of change of the participant and delivered stage-appropriate messages for management of salt consumption and procurement individually at participant’s home.

#### 2.7.2. Group counseling

In GC, all the messages were grouped according to stages of change of the participants. However, these messages were not as comprehensive as the messages included in IC. Group counseling took place in a group of 5-7 participants, and all the messages for all the stages were spoken for full audience at an establishment of community gathering.

#### 2.7.3. Mixed counseling

Mixed counseling consisted of first session of GC where all the messages of all stages of readiness of salt reduction were vocalized in a group of 5-7 participants at an establishment of community gathering. This was followed by two sessions of IC at participants’ homes consisting of delivery of stage-appropriate messages. This was again followed by one group-based session and subsequently one individual session, in the manner described previously.

#### 2.7.4. Standard care

Training of ASHAs for counseling NCD patients to adopt healthy behaviors and discarding unhealthy behaviors has been conducted by state government. ASHAs were requested to conduct ascribed visits and/or deliver ascribed messages to participants as per government mandated schedule. To avoid dissatisfaction, ASHAs in SC arm were provided with the government designed NCD module, printed and bound to appear like intervention modules.

### 2.8. Study Outcomes

Primary outcomes of the study include reduction in per capita daily salt consumption and monthly household salt procurement. Dietary salt consumption was estimated by measurement of urinary salt/sodium excretion estimated through spot urine sample and Kawasaki formula.[31] We considered thirty (30) days to be one month. For the measurement of monthly household salt procurement, we recorded dates of last two salt purchases and the amount (weight in kg) of salt purchases on these two dates. Then unitary method was used to calculate salt procured for one day at household level and salt procured for one month (30 days) was estimated.

Secondary outcomes of the trial include absolute and percentage change in SBP and change in DBP.

The following table shows the schedule of the activities of the trial.

### 2.9. Data collection

*Anganwadi* Workers (AWWs) is a cadre of CHWs working under the Integrated Child Development Services (ICDS) Scheme, catering primarily to mother and child health services.[32] AWWs of the clusters included in the intervention were approached for incentivized data collection. An incentive of INR 25.00 per participant was offered to these CHWs. KoboToolbox was used for paperless data collection.[33] Baseline data collection consisted of following sections: socio-demography of the participants (including awareness about status of hypertension and medication), salt-related knowledge (regarding daily consumption limit, health effects), knowledge of hypertension and its effects on health, cooking-related practices, purchase-related practices and self-efficacy in food purchase and consumption, and behaviors and practices related to dietary salt.

Measurement of BP was conducted in-facility at the *Anganwadi* center (AWC) or HWC (AWC is a facility for dissemination of ICDS scheme services, and AWWs are trained in measurement of BP). Measurement of BP was done using an OMRON HEM 7121 automated blood pressure monitor (OMRON, Kyoto, Japan). Measurements were conducted in clinical settings, obtaining three readings at an interval of five minutes each, and averaging the lower two readings.

### 2.10. Analysis

Summary statistics will be presented for describing the total sample and trial arms, separately. It will include measures of central tendency and dispersion of all the parameters.

An intention-to-treat analysis will be conducted after collection of end-line data of the study. Within- and between-arm comparisons will be conducted for all the outcome variables. This shall be conducted using ANOVA and Tukey’s HSD, or Kruskal-Wallis Test and Dunn’s Test for between-arm comparisons and paired sample t-test, Wilcoxon Rank Sum or Mann-Whitney U test will be used for within-arm comparisons. Pairwise comparison with corrections to reduce False Discovery Rate (FDR) will be conducted wherever necessary. Factorial ANOVA type III will be conducted for the assessment of main effects and interactions. Confidence Intervals (95%) will be used wherever required. And a p-value of 0.0.5 or less will denote statistical significance.

All the data collected via KoboToolbox shall be exported in Microsoft Excel program of Office 365 Suite (Microsoft Corporation, USA). The final data file shall be imported in RStudio IDE for further analysis in R programming language/software. Packages including but not limited to, *tidyverse[34], gtsummary[35]* and r*markdown[36]* shall be used for analysis of data.

## 3. Results

### 3.1. Recruitment, randomization, and allocation

Cluster (ASHA) and participant recruitment commenced during the month of April-2021 and completed in August-2021. All the eligible ASHAs (Table 1. N = 19) were approached for participation. Eight (8) ASHAs were finally recruited in the study, randomized, and allocated to different intervention/control arms. Figure 1 provides the depiction of flow of clusters during the intervention and plan for intention-to-treat analysis.

A list of 160 hypertensive adults (candidates) was obtained from the records maintained at HWC. Of these, 127 participants were included in the study. Reasons for exclusion included age of <30 years (5 candidates), pregnancy (5 candidates), non-residence in the village/cluster (3 candidates), ongoing psychiatric treatment (3 candidates), and 15 candidates refused participation in the study.

Among the recruited participants, 28 (22.04%) belonged to IC arm, 33 (25.98%) to GC arm, 30 (23.62%) to MC arm and 36 (28.34%) to SC arm based on the identity of the cluster.

### 3.2. Baseline characteristics

Study participants between the ages of 30 and 80 years have been recruited for the study with a mean age of 52.08 ± 12.04 years. Of these, 57.48% of the participants are females and most of the participants belong to 51-60 years of age (37 or 29.13%). The description of these and other parameters for total sample and each arm of the trial separately has been depicted in Table 3.

**Table 2.** Schedule of activities of trial.

| Activity | Baseline | Sessions-1 to 5 | End-line |
| --- | --- | --- | --- |
| Informed consent | √ |  |  |
| Induction/sensitization session | √ |  |  |
| Anthropometric measurements | √ |  |  |
| Measurement of BP at HWC | √ |  | √ |
| Behavior and practice data collection | √ |  | √ |
| Urinary sodium/salt estimation | √ |  | √ |
| Collection of salt procurement data | √ |  | √ |
| Assessment of readiness to change | √ | √ |  |
| Delivery of intervention | √ | √ |  |

**Table 3.**
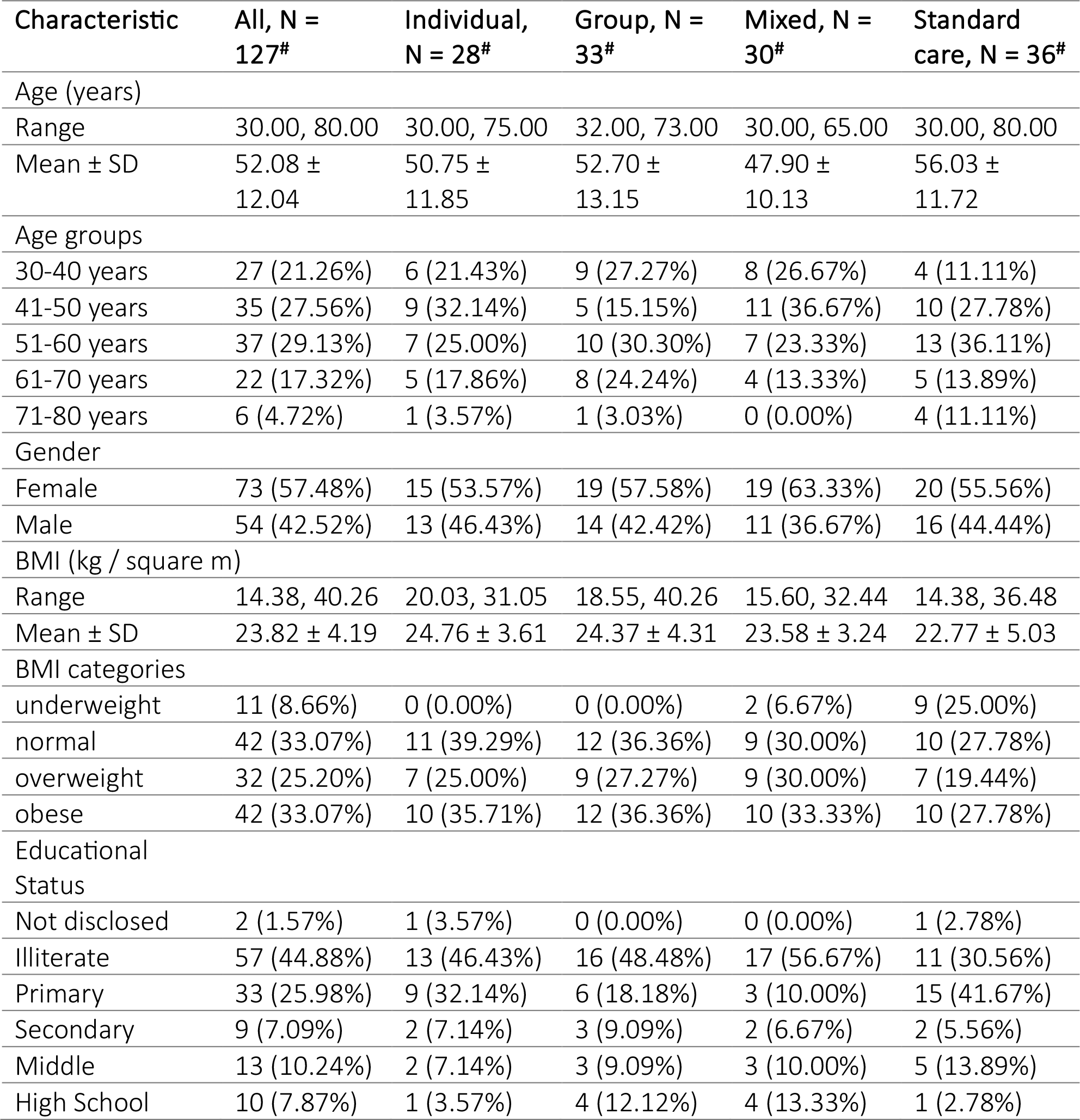
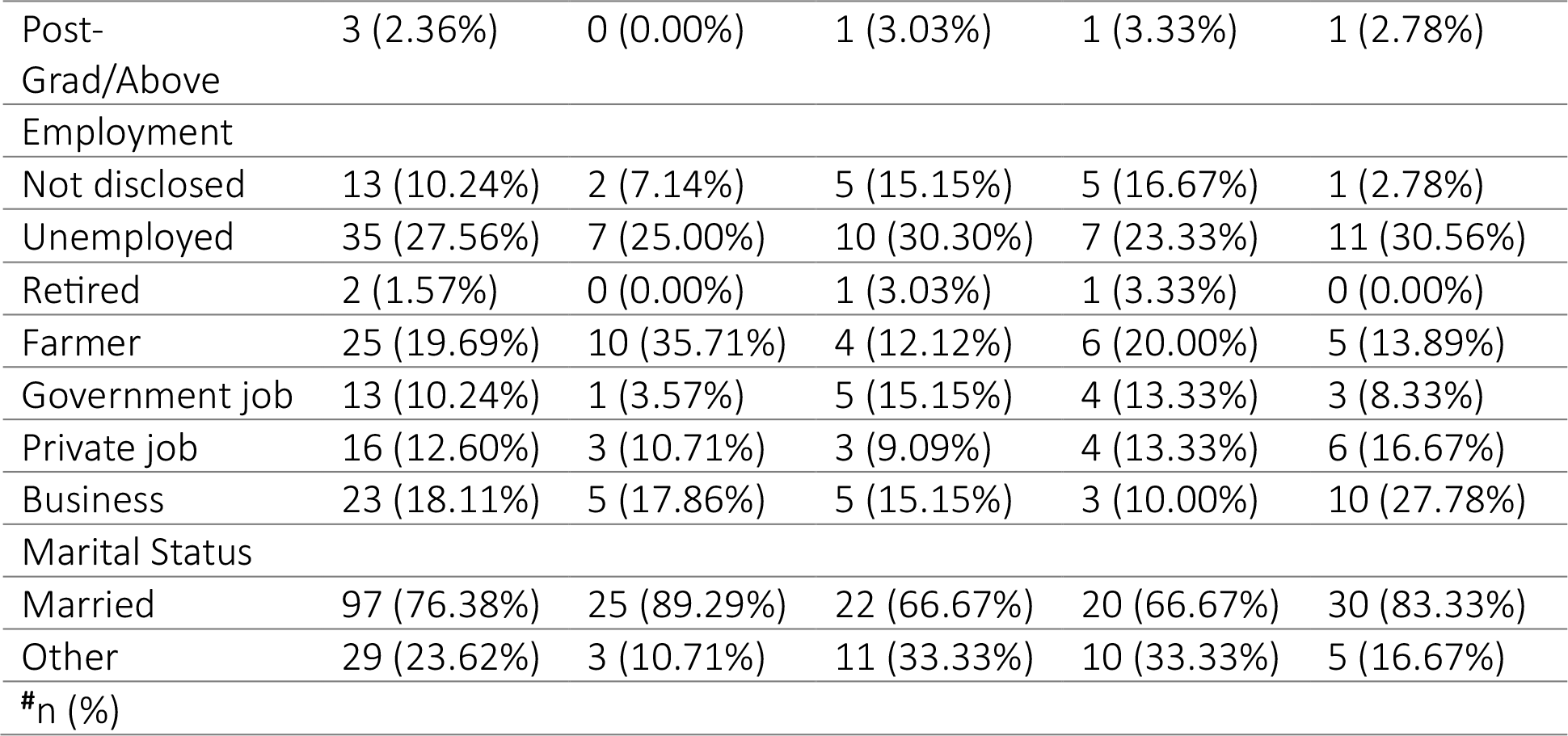
Description of the Study Participants, n = 127.

Table 4 shows the status of outcome variables at baseline. The overall per capita daily salt intake of the trial participants at baseline is observed to be 10.65 ± 2.10 grams and the monthly household salt procurement is observed to be 2.10 ± 0.93 kg. The mean SBP and DBP of all the trial participants and of trial arms separately were above 140 and 90 mm Hg, respectively.

**Table 4.** Status of Outcome variables at Baseline, n = 127.

| Outcome variable | All, N = 127<sup>#</sup> | Individual, N = 28<sup>#</sup> | Group, N = 33<sup>#</sup> | Mixed, N = 30<sup>#</sup> | Standard care, N = 36<sup>#</sup> |
| --- | --- | --- | --- | --- | --- |
| Salt intake (g/d) |  |  |  |  |  |
| Mean ± SD | $10.65 \pm 2.10$ | $11.16 \pm 2.43$ | $10.14 \pm 2.00$ | $10.82 \pm 2.09$ | $10.57 \pm 1.88$ |
| Salt procurement (kg/m) |  |  |  |  |  |
| Mean ± SD | $2.10 \pm 0.93$ | $2.18 \pm 1.05$ | $2.07 \pm 0.92$ | $2.27 \pm 1.03$ | $1.91 \pm 0.72$ |
| SBP (mmHg) |  |  |  |  |  |
| Mean ± SD | $144.62 \pm 20.63$ | $139.50 \pm 24.29$ | $143.91 \pm 17.62$ | $150.30 \pm 23.64$ | $144.53 \pm 16.77$ |
| DBP (mmHg) |  |  |  |  |  |
| Mean ± SD | $94.45 \pm 10.68$ | $93.96 \pm 8.23$ | $94.45 \pm 10.97$ | $97.17 \pm 12.81$ | $92.56 \pm 10.10$ |

## 4. Discussion

In this manuscript, we have described the rationale, design, and statistical analysis plan of a community-based cluster RCT of behavior change interventions aimed at reducing salt consumption and salt procurement at individual and household level. In addition, we have described the baseline characteristics and baseline status of outcome variables of a sample of 127 hypertensive adults recruited to participate in the study.

This trial is a part of a bigger study aimed at evaluating the efficiency of a culturally acceptable, patient-informed, family- and society-inclusive communication intervention customized according to the needs of beneficiaries and delivered by ASHAs.

The package of services rolled out in 2022, under the NPCDCS program’s Comprehensive Primary Health Care update enlists population-based screening of individuals aged 30 years or above at HWC.[37] Additionally, it also included health promotion via community-level targeted communication.[37] Therefore, we restricted the current interventional study to individuals of the same age bracket.

Recruitment of participants of our study was enabled by engaging CHWs for the process. Community Health Workers have been known to increase recruitment and retention of even hard-to-reach populations.[38] Recruitment for the study was also aided by the use of pre-existing records of the patients residing in the community. However, recruitment of ASHAs and study participants was delayed due to COVID-19 associated protocols and engagement of CHWs in COVID-19 vaccination drive.

The number of participants in the clusters ranged between 11 and 16. And the number of female participants was more than 50% in each arm. This female participation dominance, particularly in community-based studies has been observed in previous studies from rural India.[24] This might be attributed to CHWs being women themselves.

Although, participants were distributed almost equally based on their BMI, only two of the arms (MC and SC) had participants had underweight participants, of which SC had a disproportionately higher number of underweight participants (25%). The association between hypertension and BMI has been established previously.[39] We may observe an influence of weight on control of blood pressure up on daily salt consumption reduction. In our intervention, we will deliver messages pertaining to medication, particularly threats of reducing dosage or frequency of medicines, or discontinuation of treatment without medical supervision. Additionally, we have excluded participants who have altered physiological needs, such as pregnant/lactating women. Hence, our intervention poses no novel health risks to the participants. Also, we do not speculate any out-of-pocket expenditure to be borne by the participants.

The delivery of intervention is diffused across 56 days or approximately two months. The temporal distance between consecutive session increases progressively (7 days, 14 days, 28 days, and 56 days). These intervals shall enable assessment of readiness to adopt salt-restricting behaviors, and how, if at all, the readiness changes with increment in duration of reminders.

Furthermore, the study shall also attempt to generate dissecting evidence regarding comparative effectiveness of individual and group interventions in a community-based setting.

## 5. Conclusion

Our trial will report effectiveness of participant-accepted and culturally adapted behavior change communication intervention delivered via individual and group modes of counseling, as compared to standard care and to each other. Additionally, the trial will also resolve the interaction, if any, between the two modes of counseling. The results of the trial will be expandable to all hypertensive individuals above the age of 30 years, particularly to the candidates residing in rural India and/or with similar dietary practices and cultural background. The results will also be applicable to policymaking and NCD control programs.

## Data Availability

All data produced in the present study are available upon reasonable request to the authors.

## Ethics Statement

This trial was a part of a larger study undertaken by RSN for fulfilling the requirement of doctoral degree in the institution of affiliation. The ethics clearance was obtained from Intuitional Human Ethics Committee for Post Graduate Research (IHEC-PGR; reference: IHECPGRPDG067) and the trial was registered with Clinical Trial Registry of India (CTRI; CTRI/2020/11/029341).

## Supplementary data

**Supplementary Figure 1.**
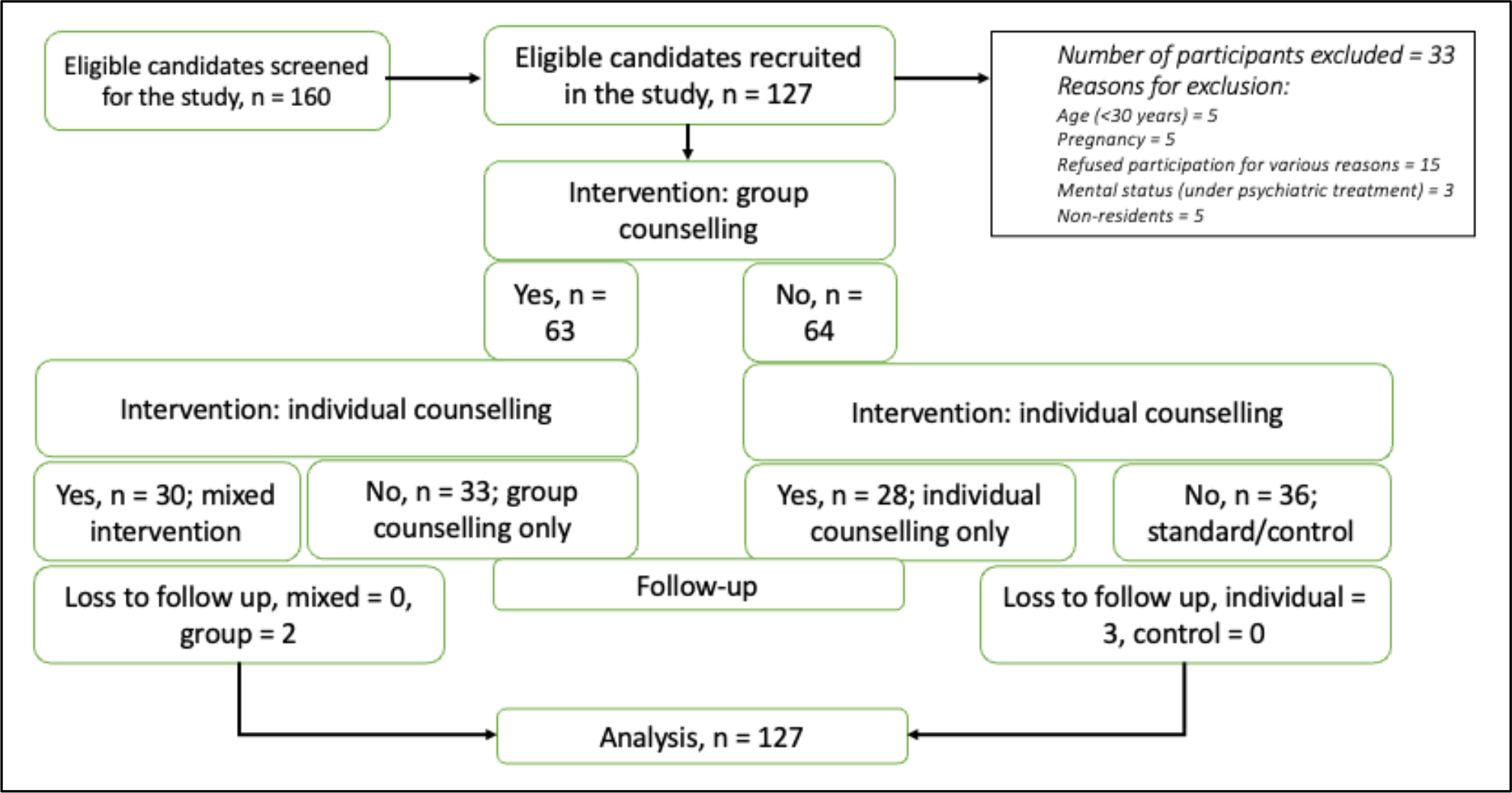
CONSORT flow diagram of the participants.

**Supplementary Table 1.** Messages included in the intervention.

| Domain | Target audience |
| --- | --- |
| Knowledge |  |
| ☐ Salt and its effects of on health | Individual and household |
| ☐ Salt reduction and its benefits | Individual and household |
| Action |  |
| ☐ Reduction of salt intake |  |
| ☐ Addition practices | Hypertensive individuals |
| ☐ Cooking practices | Cooks of the household |
| ☐ Reduction of salt procurement | Decision-makers of the household |
| Support | Family members of hypertensive individual |
| Motivation | Family members of hypertensive individual |

